# Number of ICD-10 diagnosis fields required to capture sepsis in administrative data and truncation bias: A nationwide prospective registry study

**DOI:** 10.1101/2024.07.03.24309876

**Authors:** Nina Vibeche Skei, Jan Kristian Damås, Lise Tuset Gustad

## Abstract

**Background:** In observational studies that uses administrative data, it is essential to report technical details such as the number of International Classification of Disease (ICD) coding fields extracted. This information is crucial for ensuring comparability between studies and for avoiding truncation bias in estimates, particularly for complex conditions like sepsis. Specific sepsis codes (explicit sepsis) is suggested identified by extracting 15 diagnosis fields, while for implicit sepsis, comprising an infection code combined with an acute organ failure, the number of diagnosis field remains unknown.

**Objective:** The objective was to explore the necessary number of diagnosis fields to capture explicit and implicit sepsis.

**Materials and methods:** We conducted a study utilizing The Norwegian Patient Register (NPR), which encompasses all medical ICD-10 codes from specialized health services in Norway. Data was extracted for all adult patients with hospital admissions registered under explicit and implicit sepsis codes from all Norwegian hospitals between 2008 through 2021.

**Results:** In 317,705 sepsis admissions, we observed that 105,499 ICD-10 codes were identified for explicit sepsis, while implicit sepsis was identified through 270,346 codes for infection in combination with 240,586 codes for acute organ failure. Through our analysis, we found that 55.3%, 37.0%, and 10.0% of the explicit, infection, and acute organ failure codes, respectively, were documented as the main diagnosis. The proportion of explicit and infection codes peaked in main diagnosis field, while for acute organ failure codes this was true in the third diagnosis field. Notably, the cumulative proportion reached 99% in diagnosis field 11 for explicit codes and in diagnosis field 14 for implicit codes.

**Conclusion:** Expanding the utilization of multiple diagnosis fields can enhance the comparability of data in epidemiological studies, both internationally and within countries. To make truncation bias visible, reporting guidelines should specify the number of diagnosis fields when extracting ICD-10 codes.

## Introduction

International Classification of Disease (ICD) codes are used to describe patients’ clinical characteristics and outcomes in hospital records, and these are often abstracted for research purposes [1]. Extraction of sepsis ICD codes involves specific sepsis codes (explicit sepsis) and implicit sepsis code strategy [2, 3]. The latter consist of a combination of two codes, i.e. a code for infection and a code for acute organ dysfunction. Thus, sepsis extraction strategy must involve both main condition and secondary diagnosis fields.

Previous research has revealed that increasing number of secondary diagnosis fields for explicit sepsis codes increases the accuracy of measurement, and that certain sepsis conditions are particularly prone to truncation bias if one does not examine six or more secondary diagnosis codes [4]. However, recommendations for how many secondary diagnosis fields that are needed to capture implicit sepsis are sparse.

Therefore, the aim of this study was to assess the number of diagnosis field needed to capture explicit and implicit sepsis.

## Materials and methods

We conducted a prospective descriptive registry study using The Norwegian Patient Register (NPR) that include all medical ICD-10 codes from specialized health services in Norway and has a good coverage [5, 6]. In NPR, an unlimited number of diagnosis can be reported [7], however, due privacy considerations and protection, we extracted ICD-10 codes from the main condition, the main joined condition, and up to 19 other conditions (secondary diagnosis field).

Sepsis was identified and classified using ICD-10 codes based on the Sepsis-3 definition [8]. The extraction was performed at the individual level for all patients over 18 years old with hospital admissions registered using ICD-10 codes for infection in combination with acute organ failure (implicit sepsis) and specific sepsis codes (explicit sepsis) from all Norwegian hospitals during the period from January 1, 2008, to December 31, 2021. Clinical sepsis codes (R-codes; e.g. R57.2 septic shock) is only valid in national guidelines in combination with other codes (e.g. infection code)[6], thus the R-codes were included in acute organ failure category. Infection, acute organ failure, and explicit sepsis codes were classified as binary variables (0 and 1), i.e. either absent or present. These codes were retrieved from the main diagnosis fields, main joined diagnosis field, and 19 secondary diagnosis fields. As the percentage of ICD-10 codes in the joined diagnosis field to main diagnosis was only 0.1% in each category, we merged the joined diagnosis field with main diagnosis and named the diagnosis field 1. The secondary diagnosis fields were denoted as diagnosis field 2 through 20. Details on the ICD-10 codes extracted are priory published [3].

## Statistical Analysis

For every diagnosis field, we counted the number of explicit sepsis, infection, and acute organ failure ICD-10 codes and calculated the proportion by dividing the number of each code by the total number of corresponding diagnoses. We then reported the proportion of codes per group (explicit, infection, or acute organ failure) for each diagnosis field, as well as the cumulative proportion. We used the Stata software package (version 16, Stata Corp, TX, USA) for all statistical analyses.

## Ethics

The study was approved by the Regional Committee for Medical and Health Research Ethics in Eastern Norway (2019/42772) and the Data Access Committee in Health North-Trøndelag (2021/184). The analyses were conducted in the Services for Sensitive Data at the University of Oslo.

## Results

Among 12.6 million discharges from 2008 through 2021, a total of 317 705 patients was identified with more than one ICD-10 sepsis code. Of these, 105 499 ICD-10 codes were identified for explicit sepsis, while implicit sepsis was identified through 270 346 codes for infection in combination with 240 586 codes for acute organ failure (Table 1).

**Table 1.**
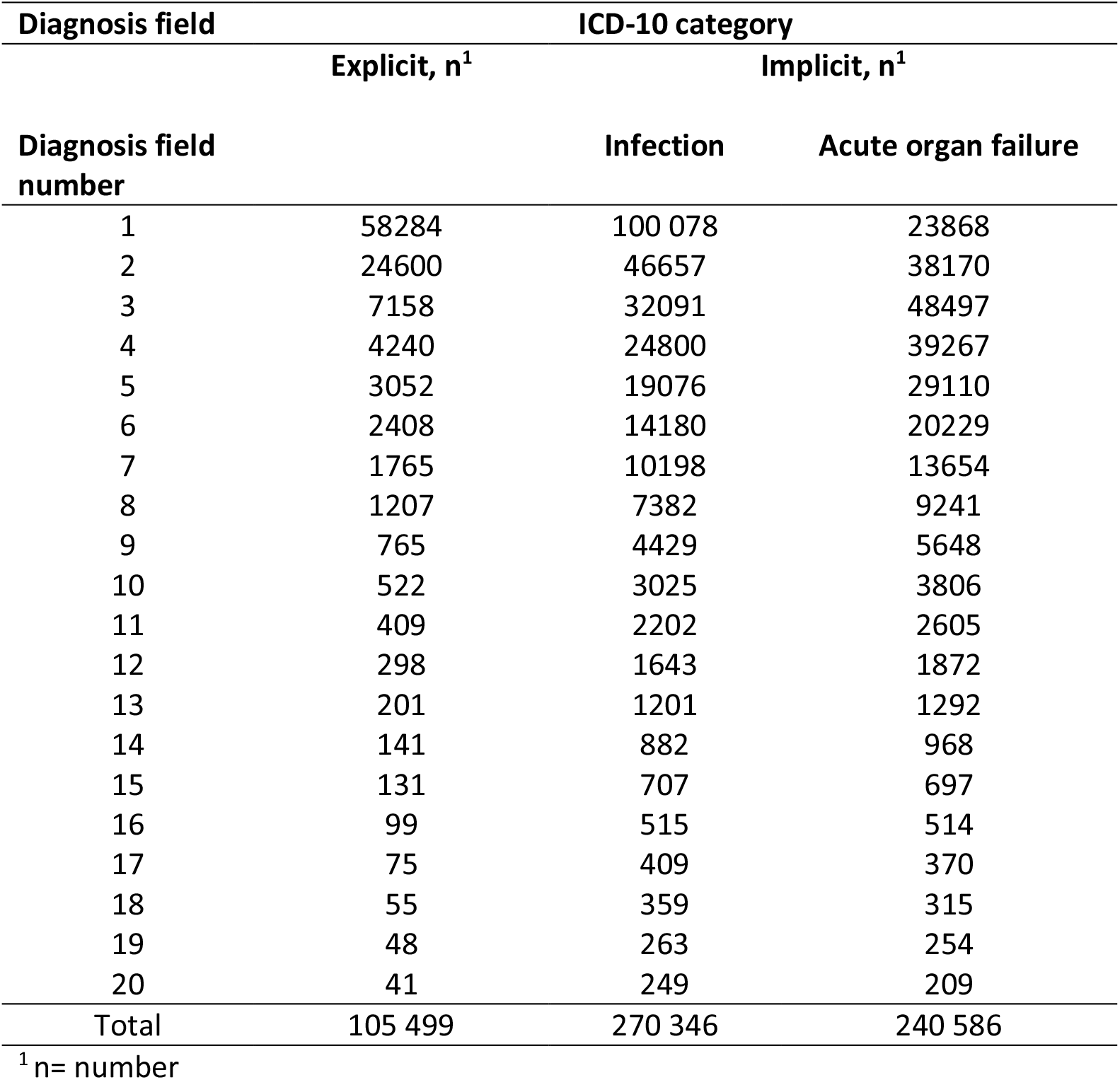
Number of ICD-10 sepsis codes in main and 19 secondary diagnosis fields.

| Diagnosis field | ICD-10 category |  |  |
| --- | --- | --- | --- |
|  | Explicit, n<sup>1</sup> | Implicit, n<sup>1</sup> |  |
| Diagnosis field number |  | Infection | Acute organ failure |
| 1 | 58284 | 100 078 | 23868 |
| 2 | 24600 | 46657 | 38170 |
| 3 | 7158 | 32091 | 48497 |
| 4 | 4240 | 24800 | 39267 |
| 5 | 3052 | 19076 | 29110 |
| 6 | 2408 | 14180 | 20229 |
| 7 | 1765 | 10198 | 13654 |
| 8 | 1207 | 7382 | 9241 |
| 9 | 765 | 4429 | 5648 |
| 10 | 522 | 3025 | 3806 |
| 11 | 409 | 2202 | 2605 |
| 12 | 298 | 1643 | 1872 |
| 13 | 201 | 1201 | 1292 |
| 14 | 141 | 882 | 968 |
| 15 | 131 | 707 | 697 |
| 16 | 99 | 515 | 514 |
| 17 | 75 | 409 | 370 |
| 18 | 55 | 359 | 315 |
| 19 | 48 | 263 | 254 |
| 20 | 41 | 249 | 209 |
| Total | 105 499 | 270 346 | 240 586 |

<sup>1</sup> n= number

The main condition was recorded in 55.3% of the explicit codes, 37.0% of the infection codes and 10.0% of the acute organ dysfunction codes. The proportion of explicit and infection codes peaked in main diagnosis field, while for acute organ failure codes this was true in the third diagnosis field (Fig 1). Over 99% of the explicit sepsis cases were covered by diagnosis field 11, and the same yield for infection and acute organ failure codes in diagnosis field 14 (Fig 2).

**Fig 1.**
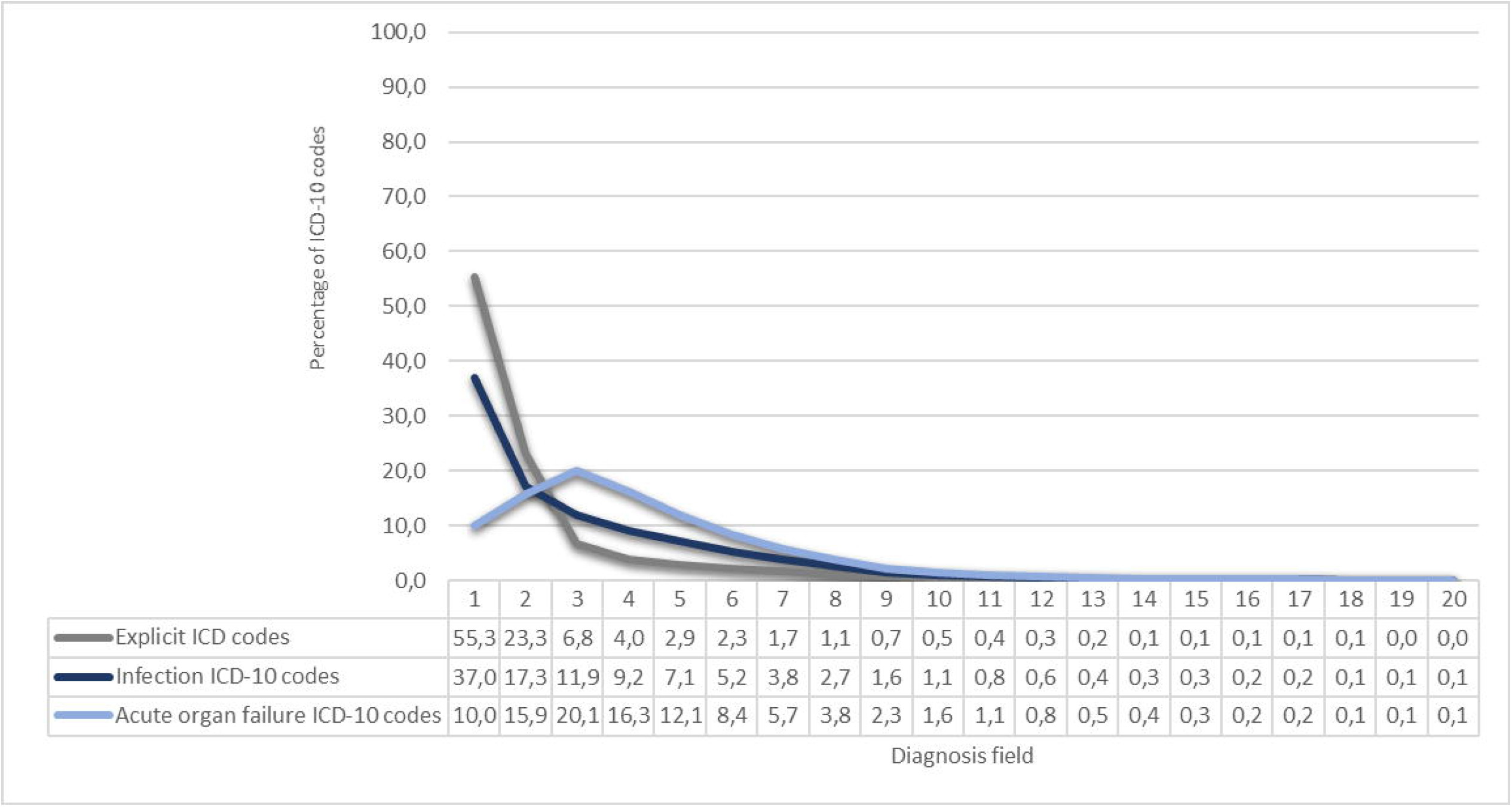
Number and (%) of ICD-10 codes in diagnoses field one through 20 for explicit sepsis, infection and organ dysfunction sepsis codes.

**Fig 2.**
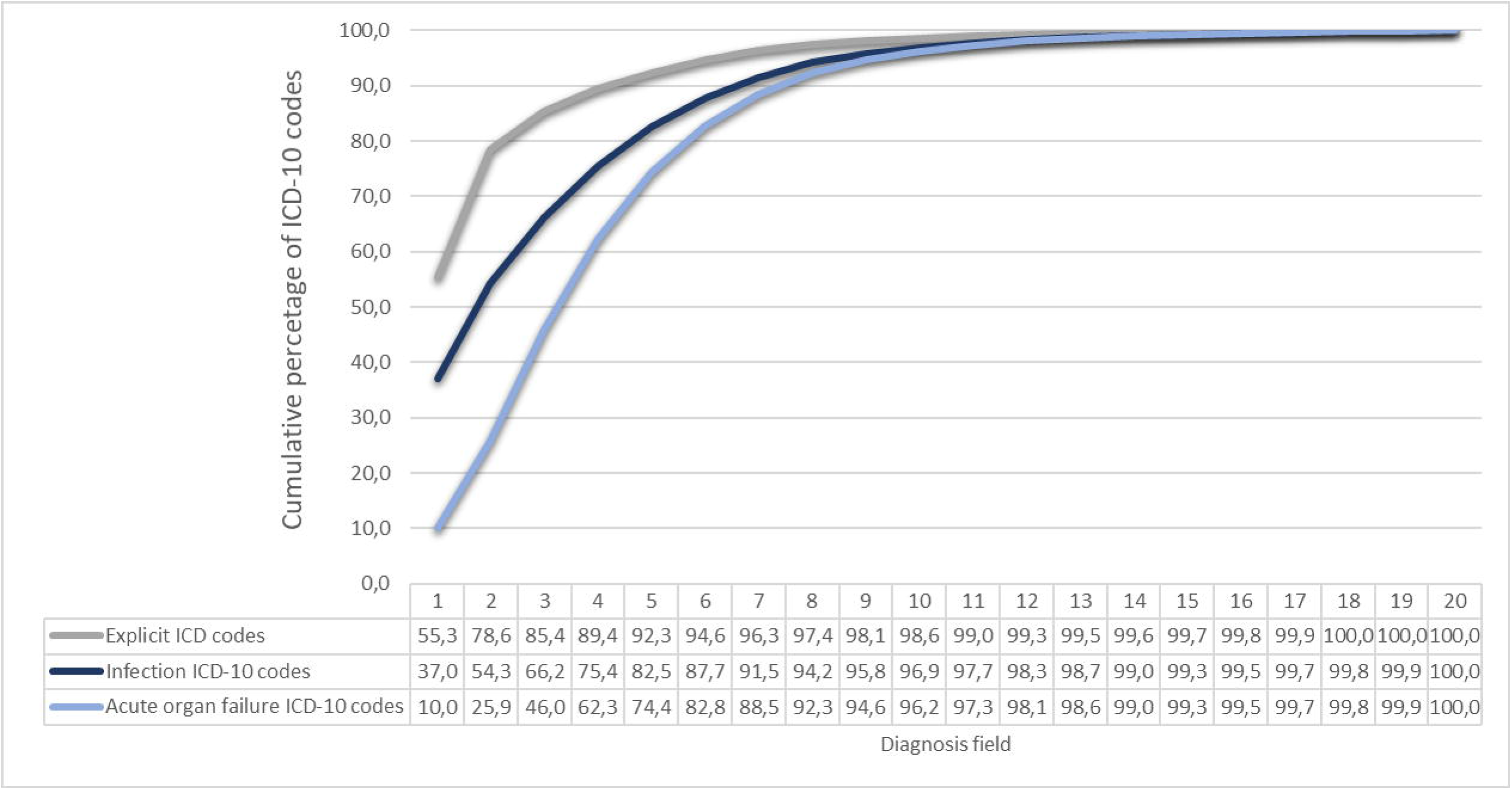
The cumulative percentage of ICD-10 codes for explicit sepsis, infection and organ dysfunction sepsis codes by diagnosis field 1 through 20.

## Discussion

In this nationwide article, we present proportions of explicit and implicit codes as main condition and up to 19 secondary diagnoses. Our findings reveals that the majority of explicit codes (55%) was listed as main condition, while this yield for only 37% of the infection codes and for 10% of the acute organ failure codes. Notably, cumulative proportion reached 99% in diagnosis field 11 and 14 for explicit and implicit codes, respectively.

To our knowledge, no previous study has examined the necessary medical diagnosis fields to extract implicit sepsis. Our findings of how many diagnosis fields requited to capture explicit sepsis are somewhat lower than a previous study commissioned by the World Health Organization investigating postoperative explicit sepsis codes among twenty countries suggesting that at least 15 secondary diagnosis fields is optimal for relevant clinical information using ICD-10 codes [4]. Unlike postoperative sepsis, it is probable that a non-postoperative sepsis admission (e.g. acute sepsis) will be classified as a primary or at least as an early secondary diagnosis. Therefore, our wider inclusion of specific sepsis codes may account for the differing outcome.

Information about the number diagnosis fields used during extraction of data is missing in reporting guidelines for observational studies [9]. One of the challenges when comparing studies is the differences in national ICD-10 coding guidelines. A prior sepsis ICD coding validation study of 22 international studies on a population level compared five strategies [10]. They found that R-codes and explicit sepsis coding strategies may underestimate sepsis incidence by 3.5-fold and 3-fold, respectively. However, in many of these epidemiological studies of sepsis, information about the technical extraction strategy involving the number of diagnosis fields is missing, making it difficult to compare national sepsis incidence. Our study has revealed that extracting sepsis codes of less than 11 diagnosis fields for explicit and 14 fields for implicit sepsis may introduce a truncation bias, potentially leading to underestimation of incidence.

Our study boasts several notable strengths. Firstly, it draws on data from all public hospitals in Norway spanning 14 years. Secondly, the comprehensive assessment of diagnosis fields used to identify sepsis, covering implicit and explicit sepsis codes, adds to the robustness of our findings. Thirdly, in Norway, reporting ICD-10 codes to NPR is obligatory and undergoes quality checks conducted by the National Service of Validation and completeness analysis. This ensures that our extraction of ICD-10 codes has minimal missing, incomplete, or unknown discharge codes [7]. Furthermore, the extraction of medical codes for sepsis identification previous used by other researchers further strengthens the integrity of our study [2, 11, 12]. Lastly, in contrast to many other countries, available numbers of secondary diagnosis fields in the data set to capture events are unlimited in Norway [4]. However, in our study we extracted ICD-10 codes from 19 secondary diagnosis fields due to data minimization. Therefore, we cannot rule out that extraction from more diagnosis fields could have increased the diagnosis fields needed to capture sepsis.

## Conclusion

This nationwide study suggest that at least 11 diagnosis fields are required in extraction of explicit sepsis codes and 14 diagnosis fields to extract implicit sepsis codes. Increasing the use of multiple diagnosis fields will improve international and intra-national comparability of data in epidemiological studies. Additionally, it will enhance the quality of analyses, allowing for better utilization of results. Reporting guidelines should include how many diagnosis fields used when extracting ICD-10 codes from administrative data in order to make truncation bias visible.

## Data Availability

No additional data available. We do not have ethical approval to deposit our datasets in publicly available repositories. Researchers need approval by the Regional Ethical Committee for handling of NPR data files. The NPR has precise information on all data exported to different projects and there are no restrictions regarding data export given Regional Etical comitee approval.

## Acknowledgement

None

## References

1. Quan H, Moskal L, Forster AJ, Brien S, Walker R, Romano PS, et al. International variation in the definition of ‘main condition’ in ICD-coded health data. Int J Qual Health Care. 2014;26(5):511–5.

2. Rudd KE, Johnson SC, Agesa KM, Shackelford KA, Tsoi D, Kievlan DR, et al. Global, regional, and national sepsis incidence and mortality, 1990-2017: analysis for the Global Burden of Disease Study. Lancet. 2020;395(10219):200–11.

3. Skei NV, Nilsen TIL, Mohus RM, Prescott HC, Lydersen S, Solligård E, et al. Trends in mortality after a sepsis hospitalization: a nationwide prospective registry study from 2008 to 2021. Infection. 2023.

4. Drösler SE, Romano PS, Sundararajan V, Burnand B, Colin C, Pincus H, Ghali W. How many diagnosis fields are needed to capture safety events in administrative data? Findings and recommendations from the WHO ICD-11 Topic Advisory Group on Quality and Safety. Int J Qual Health Care. 2014;26(1):16–25.

5. Norwegian Patient Registry [Available from: https://www.helsedirektoratet.no/english.

6. ICD-10 og ICD-11: Directorate of e-health; 2022 [updated April 2022. Available from: https://www.ehelse.no/kodeverk-og-terminologi/ICD-10-og-ICD-11.

7. Bakken IJ, Ariansen AMS, Knudsen GP, Johansen KI, Vollset SE. The Norwegian Patient Registry and the Norwegian Registry for Primary Health Care: Research potential of two nationwide health-care registries. Scand J Public Health. 2020;48(1):49–55.

8. Singer M, Deutschman CS, Seymour CW, Shankar-Hari M, Annane D, Bauer M, et al. The Third International Consensus Definitions for Sepsis and Septic Shock (Sepsis-3). JAMA. 2016;315(8):801–10.

9. network e. Enhancing the QUAlity and Transparency Of health Research 2024 [cited 2024 June 26]. Available from: https://www.equator-network.org/reporting-guidelines/strobe/.

10. Fleischmann-Struzek C, Thomas-Ruddel DO, Schettler A, Schwarzkopf D, Stacke A, Seymour CW, et al. Comparing the validity of different ICD coding abstraction strategies for sepsis case identification in German claims data. PLoS One. 2018;13(7):e0198847.

11. Angus DC, Linde-Zwirble WT, Lidicker J, Clermont G, Carcillo J, Pinsky MR. Epidemiology of severe sepsis in the United States: analysis of incidence, outcome, and associated costs of care. Crit Care Med. 2001;29(7):1303–10.

12. Martin GS, Mannino DM, Eaton S, Moss M. The Epidemiology of Sepsis in the United States from 1979 through 2000. New England Journal of Medicine. 2003;348(16):1546–54.

